# Identify Patients at Risk of HIV Using a Clinical Large Language Model from Electronic Health Records

**DOI:** 10.64898/2026.04.21.26351427

**Authors:** Yiyang Liu, Ziyi Chen, Suman Pogul, Hwayoung Cho, Mattia Prosperi, Yonghui Wu

## Abstract

This study developed a large language model (LLM)-based solution to identify people at HIV risk using electronic health records. We transformed structured EHR data, including demographics, diagnoses, and medications, into narrative descriptions ordered by visit date and applied GatorTron, a widely used clinical LLM trained on 82 billion words of de-identified clinical text. We compared GatorTron with traditional machine learning models, including LASSO and XGBoost. We identified a cohort with 54,265 individuals, where only 3,342 (6%) had new HIV diagnoses. Our LLM solution, based on GatorTron, achieved excellent performance, reaching an F1 score of 53.5% and an AUC of 0.88, comparable to traditional machine learning approaches. Subgroup analysis showed that, across age, sex, and race/ethnicity groups, both LLM and traditional models achieved AUCs above 0.82. Interpretability analyses showed broadly consistent patterns across LLM models and traditional machine learning models.

## Introduction

HIV infection brings an enormous burden to healthcare systems worldwide. Over 40 million people globally and around 1.2 million people in the United Stated living with HIV [1, 2]. There is growing interest in leveraging electronic health records (EHRs) to identify patients with HIV risk to counter this worldwide threat [3]. Several computational models have been developed and evaluated across a variety of EHR data sources, ranging from a single health system [4-8] to multi-health-system clinical networks [9-12], within the United States [4-8, 10, 13, 14] and abroad [9, 12]. Previous studies have developed various computational approaches, such as logistic regression[13] and traditional machine learning methods, examined a wide range of socio-demographic and clinical predictors [4-10, 12, 14], and reported promising performance from different populations, including male-dominated [5, 6, 13], women-specific [4, 7, 11], and emerging adults samples [10]. These models are promising in helping providers identify individuals at elevated risk of HIV and initiate risk-reduction conversations [15], recommend HIV prevention services such as pre-exposure prophylaxis (PrEP) [16, 17] and frequent HIV testing [12].

Nevertheless, existing models based on traditional machine learning models and EHRs are known to suffer from the feature sparsity caused by the complex medical coding systems. For example, the International Classification of Diseases (ICD) codes are mainly proposed for billing purposes, and there are over 60,000 individual disease codes and multiple versions, such as ICD-9 and ICD-10. When directly using the original diagnosis codes as predictors for a real-world HIV cohort, there are often limited subjects for each diagnosis code, causing a sparsity issue that diminishes prediction power. Therefore, researchers often have to carefully select the predictors based on domain knowledge [5, 11, 18]. Even with data-driven methods [4], such as frequency filters [11], there is still a need to group them into broader categories, such as the PeWAS disease group, to alleviate the sparsity issue. Preprocessing medication use can be even more challenging, as a single condition may be treated with multiple regimens, and medications often exist in both generic and branded forms with varying dosages and packaging. These variations correspond to different normalized names for clinical drugs (RxNorm) and national drug code (NDC), resulting in a single medication being associated with multiple identifiers.

Large language models (LLMs) offer a convenient way to develop clinical prediction models by using narrative descriptions. Trained with massive amounts of text, LLMs have demonstrated advanced capabilities not only in natural language processing (NLP) [19] but also in disease risk prediction [20]. Instead of manually coded one-hot encoding, researchers can convert structured EHR codes into text representations, which allows LLMs to assess disease risk according to the semantics carried in the medical text. Bidirectional Encoder Representations from Transformers (BERT)-based LLMs [21] are designed to capture contextual meaning in both directions of text and are particularly well suited for disease prediction. Building on this architecture, our team developed GatorTron [22, 23], an encoder-based LLM with up to 8.9 billion parameters trained on 90 billion words of medical text. Past work demonstrated that GatorTron prediction overperformed traditional machine learning and deep learning methods in predicting heart failure risk among cancer patients using structured EHR [20]. In this work, we extend the application of GatorTron to HIV risk prediction. Specifically, we compare its performance with traditional machine learning methods and assess model fairness across demographic groups and explore model interpretability.

## Methods

### 2.1 Data source and study sample

We used longitudinal EHR data from 2012 to 2023 obtained from the OneFlorida+ Data Consortium [24]. It includes claims-linked EHRs for over 20 million patients from multiple healthcare systems across Florida, Georgia, and Alabama. All data were standardized to the Patient-Centered Outcomes Research Institute (PCORI) Common Data Model (PCORnet CDM [25] and released as a Health Insurance Portability and Accountability Act (HIPAA)-compliant limited dataset.

We identified an HIV cohort using the same inclusion criteria that have been verified in our prior HIV risk prediction study among women [11]. Briefly, we used a case cohort study design with the following inclusion criteria: 1) age 13 years or older, 2) having at least one year of EHR records, 3) one year of continuous insurance enrollment, and 4) three clinical encounters. The index date was defined as January 1 of the year of HIV diagnosis for cases. For controls, the index date was January 1 of a randomly selected year in which they met the inclusion criteria, with each individual included only once in the cohort. The outcome was a new HIV diagnosis in the calendar year following the index date. EHR data documented before the index date were used for predictor extraction.

#### 2.2.1 Data preprocessing for traditional machine learning models

Demographic variables included sex (female or male), age at the index date, and race/ethnicity, which was categorized as Hispanic, non-Hispanic White, non-Hispanic Black, or other. Primary insurance payer was extracted and grouped into Medicare, Medicaid, private insurance, other government, self-pay/no insurance, and other/unknown.

Clinical features were identified based on domain knowledge and prior literature on HIV risk and supplemented with additional data-driven features. The comprehensive list of expert-selected features and their corresponding ICD-9 and ICD-10 codes was provided in a prior publication focused on HIV risk prediction among women [11]. Briefly, these predictors included screening and counseling related to sexual or reproductive health, diagnoses and laboratory results for various sexually transmitted infections (STIs), diagnoses of mental health and substance use disorders, and diagnoses of chronic conditions included in the Charlson Comorbidity Index [26]. In addition, we applied a data-driven feature selection procedure to extract additional clinical features based on diagnostic code frequency. All ICD-9 and ICD-10 codes were hierarchically mapped to PheCodes[27], and PheCodes with a frequency greater than 3% were included.

All clinical features were one-hot encoded across two temporal windows: 1) in the past 12 months, and 2) in the lifetime record but not in the past 12 months. Finally, all features were examined for multicollinearity, and features with high collinearity were excluded from further analysis.

#### 2.2.2 Data preprocessing for large language models

All socio-demographic variables were converted into narrative text. These included the same variables as described above, with the addition of gender identity. For clinical features, all ICD-9 and ICD-10 diagnosis codes were hierarchically mapped to PheCodes, and the corresponding PheCode text descriptions were extracted. For ICD codes that could not be mapped to a PheCode, the raw text descriptions of the ICD codes were used instead. Unlike the traditional machine learning models, which applied frequency-based feature filtering, all diagnoses were included and converted into text descriptions. Prescription data were processed similarly. The raw EHR data contained RxNorm Concept Unique Identifiers (RxCUIs) in the prescription table. We utilized APIs provided by the U.S. National Library of Medicine [28] to map all RxCUIs to their corresponding active ingredient names and convert these into text.

For each patient, converted text data from all encounters documented before the index date were concatenated in chronological order based on encounter dates. Due to LLM input token limitations, fewer than 7.8% of patients with a large number of encounters or diagnoses had their text truncated after 512 tokens. A comparison of the data preprocessing steps for traditional machine learning and LLM models is presented in Figure 1.

**Figure 1.**
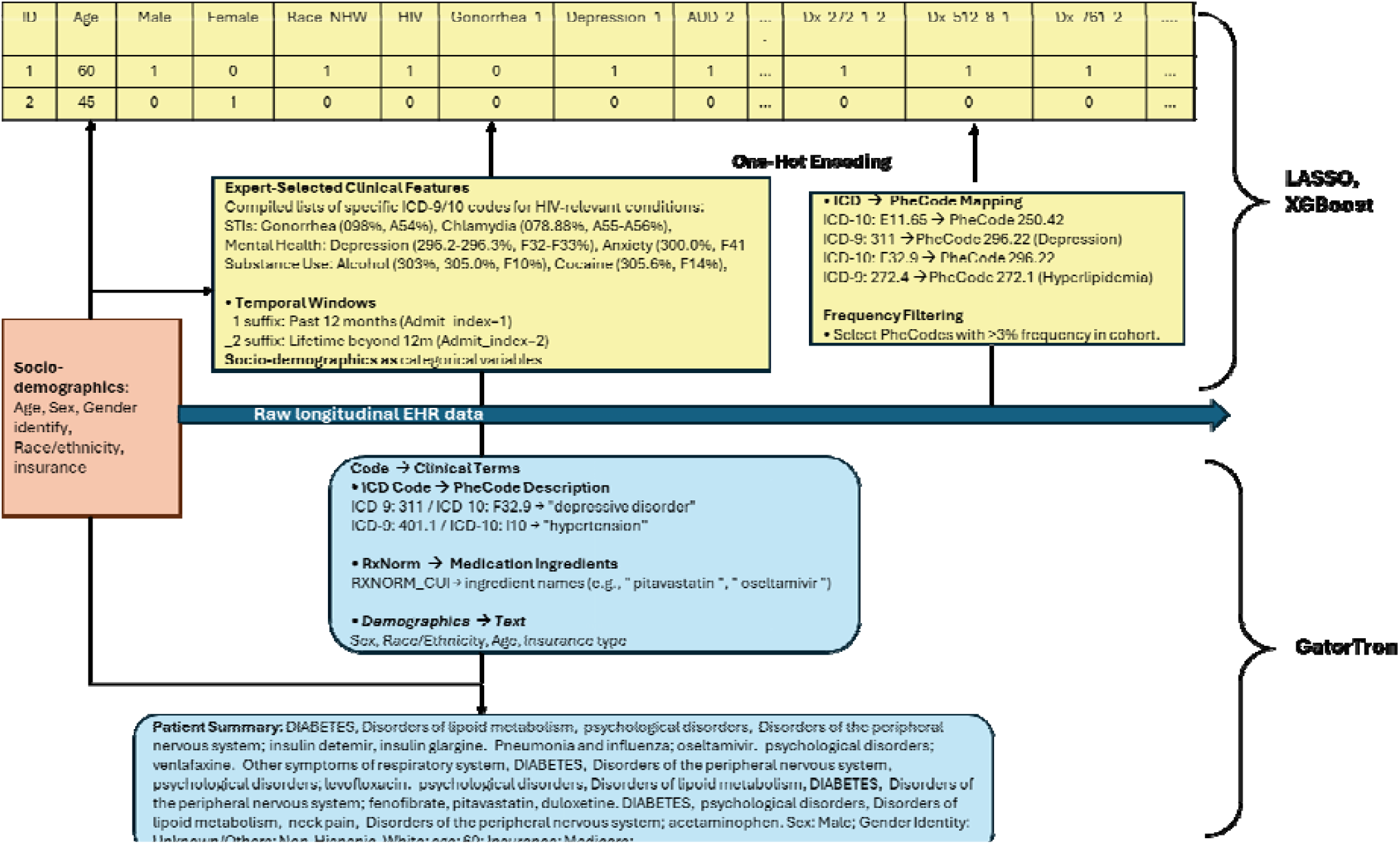
Data preprocessing steps for traditional machine learning and for GatorTron LLM

### 2.3 Machine learning models

#### Large language models

Large language models. We randomly split the dataset into training, validation, and test sets with a 6:2:2 ratio, resulting in 32,559, 10,853, and 10,853 samples in each subset, respectively. For LLM, we fine-tuned the pre-trained GatorTron-345m model, which was pretrained using a dataset of around 82 billion words of de-identified clinical notes, 6.1 billion words from PubMed CC0, and 2.5 billion words from WikiText. Fine-tuning was performed for 20 epochs with a learning rate of 1e-5. We used a maximum sequence length of 512 tokens with lowercased input text. A warm-up strategy was applied using a warm-up ratio of 0.1, followed by learning rate decay. A dropout rate of 0.1 was used to mitigate overfitting. During training, we monitored performance on the validation set to select the best model checkpoint based on the F1 score. Final performance was evaluated on the test set.

#### Traditional machine learning

For traditional machine learning, we used both least absolute shrinkage and selection operator (LASSO) logistic regression and extreme gradient boosting (XGBoost). LASSO was selected for its ease of interpretability. XGBoost, an ensemble tree-based algorithm, was included to capture potential non-linear relationships and interactions between features, and was selected based on its strong performance in predicting HIV risk among women in our prior work [11]. The training and validation sets were combined for training of both models using five-fold cross-validation, and performance was evaluated on the same test set used by LLM.

### 2.4 Model evaluation and interpretability

Model performance was evaluated on the testing dataset using six metrics: precision (positive predictive value, PPV), recall (sensitivity), specificity, F1 score, accuracy, and area under the receiver operating characteristic curve (AUC). Precision (PPV) represents the proportion of predicted positive cases that are true positives, and recall (sensitivity) represents the proportion of actual positive cases correctly identified by the model. Specificity reflects the proportion of actual negative cases correctly classified. The F1 score calculates the harmonic mean of precision and recall, balancing the trade-off between false positives and false negatives. Accuracy reflects the overall proportion of correctly classified instances. AUC reflects the trade-off between sensitivity and specificity, and represents the model’s overall ability to correctly classify positive and negative cases across all possible classification thresholds.

For the LLM models, performance was evaluated under three text input setups: (1) text extracted from diagnosis codes only (Dx only); (2) diagnoses plus socio-demographic information (Dx+Dem); and (3) diagnoses, socio-demographics, and prescription medications (Dx+Dem+Rx). For the traditional machine learning models, input features included all demographic variables as well as clinical features derived from both expert-selected and frequency-filtered methods.

To assess model fairness, we examined performance across key demographic subgroups, including sex (female, male), age group (13–29, 30–44, ≥45 years), and race/ethnicity (non-Hispanic White, non-Hispanic Black, and Hispanic).

To enhance the interpretability of the LLM, the LIME (Local Interpretable Model-agnostic Explanations) framework [29] was employed to visualize the most influential narrative features. We analyzed the attention weights assigned by the Dx+Dem+Rx fine-tuned model on the test dataset, delivering powerful insights into the core narrative elements that shape its predictions. Important subword features are highlighted in color, where orange indicates features contributing to a higher predicted HIV risk, and blue indicates features contributing to a lower predicted risk; the darker the color, the stronger the feature’s influence. For LASSO logistic regression, we reported odds ratios (ORs) with 95% confidence intervals for the top 20 features. For XGBoost, SHapley Additive exPlanations (SHAP) values were used to quantify the contributions of the top 20 features.

## Results

Our study cohort consisted of 54,265 people, including 3,342 people newly diagnosed with HIV (cases) and 50,923 people never diagnosed with HIV (non-cases). The sample characteristics are shown in Table 1. Overall, 59.6% of the sample were female, 74.4% were aged 45 or older, over half were non-Hispanic White, 23.7% had private insurance, and 16.0% had Medicare. A higher proportion of males, younger age, non-Hispanic Black, past year diagnosis of sexually transmitted infections, and substance use disorders were observed among cases relative to non-cases.

**Table 1.** Study sample characteristics, n=54,265.

| Characteristics | Total, n=54,265 | Case (new HIV diagnosis), n= 3,342 | Non-case (people without HIV), n=50,923 |
| --- | --- | --- | --- |
| Sex |  |  |  |
| Male | 21,943 (40.4%) | 1,978 (59.2%) | 19,965 (39.2%) |
| Female | 32,321 (59.6%) | 1,364 (40.8%) | 30,957 (60.8%) |
| Other | 1 (0.0%) | 0 (0.0%) | 1 (0.0%) |
| Age group |  |  |  |
| 13-29 | 3,740 (6.9%) | 688 (20.6%) | 3,052 (6.0%) |
| 30-44 | 10,131 (18.7%) | 954 (28.5%) | 9,177 (18.0%) |
| 45+ | 40,394 (74.4%) | 1,700 (50.9%) | 38,694 (76.0%) |
| Race/ethnicity |  |  |  |
| Non-Hispanic White | 28,571 (52.7%) | 835 (25.0%) | 27,736 (54.5%) |
| Non-Hispanic Black | 12,830 (23.6%) | 1,810 (54.2%) | 11,020 (21.6%) |
| Hispanic | 10,092 (18.6%) | 625 (18.7%) | 9,467 (18.6%) |
| Other | 2,772 (5.1%) | 72 (2.2%) | 2,700 (5.3%) |
| Primary insurance payer (%) |  |  |  |
| Medicare | 8,693 (16.0%) | 228 (6.8%) | 8,465 (16.6%) |
| Medicaid | 2,512 (4.6%) | 257 (7.7%) | 2,255 (4.4%) |
| Other governmental | 1,311 (2.4%) | 167 (5.0%) | 1,144 (2.2%) |
| Private | 12,879 (23.7%) | 360 (10.8%) | 12,519 (24.6%) |
| Self-pay/no insurance | 1,607 (3.0%) | 184 (5.5%) | 1,423 (2.8%) |
| Unknown/Other | 27,263 (50.2%) | 2,146 (64.2%) | 25,117 (49.3%) |
| Diagnosis in the past year |  |  |  |
| Chlamydia | 93 (0.2%) | 4 (0.1%) | 89 (0.2%) |
| Gonorrhea | 31 (0.1%) | 8 (0.2%) | 23 (0.0%) |
| Syphilis | 44 (0.1%) | 13 (0.4%) | 31 (0.1%) |
| HPV | 251 (0.5%) | 18 (0.5%) | 233 (0.5%) |
| Herpes | 217 (0.4%) | 25 (0.7%) | 192 (0.4%) |
| Trichomoniasis | 99 (0.2%) | 17 (0.5%) | 82 (0.2%) |
| Hepatitis B | 162 (0.3%) | 24 (0.7%) | 138 (0.3%) |
| Hepatitis C | 780 (1.4%) | 70 (2.1%) | 710 (1.4%) |
| Depression | 4,949 (9.1%) | 223 (6.7%) | 4,726 (9.3%) |
| Anxiety | 5,542 (10.2%) | 180 (5.4%) | 5,362 (10.5%) |
| Tobacco use disorder | 4,401 (8.1%) | 390 (11.7%) | 4,011 (7.9%) |
| Alcohol use disorder | 1,408 (2.6%) | 153 (4.6%) | 1,255 (2.5%) |
| Cannabis use disorder | 670 (1.2%) | 104 (3.1%) | 566 (1.1%) |
| Cocaine use disorder | 549 (1.0%) | 131 (3.9%) | 418 (0.8%) |
| Opioid use disorder | 603 (1.1%) | 70 (2.1%) | 533 (1.0%) |

As shown in Table 2, the LLM model demonstrated a noticeable improvement in performance after demographic factors were added to the clinical narratives based solely on diagnosis codes (e.g., AUC increased from 0.74 to 0.87, sensitivity from 19.0% to 46.7%, and F1 score from 25.2% to 53.9%). When medication information was further incorporated, only a marginal increase in AUC was observed (0.87 to 0.88), accompanied by decreases in precision (74.0% to 62.7%), Specificity (99.0 to 98.2), F1 score (53.9% to 53.5%), and accuracy (95.6% to 95.1%). Overall, the final LLM model achieved comparable discriminative performance to the traditional machine learning methods (AUC = 0.88 for LLM vs. 0.89 for XGBoost and 0.88 for LASSO). The LLM model showed higher specificity, overall accuracy and F1 score compared with the traditional machine learning methods, but lower sensitivity/recall.

**Table 2.** Comparison of model performance metrics for the clinical large language model versus traditional machine learning models.

|  | Precision (PPV) | Recall (sensitivity) | Specificity | F1 score | Accuracy | AUC score |
| --- | --- | --- | --- | --- | --- | --- |
| **LLM model** |  |  |  |  |  |  |
| Model 1: Dx only | 37.5% | 19.0% | 98.0% | 25.2% | 93.2% | 0.74 |
| Model 2: Dx+Dem | 74.0% | 42.4% | 99.0% | 53.9% | 95.6% | 0.87 |
| Model 3: Dx+Dem+Rx | 62.7% | 46.7% | 98.2% | 53.5% | 95.1% | 0.88 |
| **XGBoost model** |  |  |  |  |  |  |
| Dem+Dx (expert+freq) | 30.0% | 69.9% | 89.5% | 42.0% | 88.3% | 0.89 |
| **LASSO model** |  |  |  |  |  |  |
| Dem+Dx (expert+freq) | 26.1% | 69.8% | 87.2% | 38.0% | 86.2% | 0.87 |

When examining model performance across different demographic groups (Table 3), both the LLM and traditional machine learning methods demonstrated good to excellent discriminative performance across sex, age, and racial/ethnic groups, with AUCs ranging from 0.82 to 0.89.

**Table 3.** Subgroup analysis across different age, sex, and racial/ethnic groups.

|  | Recall (sensitivity) | Specificity | AUC score |
| --- | --- | --- | --- |
| **Sex specific** |  |  |  |
| LLM |  |  |  |
| Male | 49.3% | 97.5% | 0.88 |
| Female | 42.4% | 98.6% | 0.87 |
| XGBoost |  |  |  |
| Male | 76.5% | 83.5% | 0.88 |
| Female | 59.2% | 93.3% | 0.88 |
| LASSO |  |  |  |
| Male | 76.2% | 80.3% | 0.86 |
| Female | 59.2% | 91.7% | 0.86 |
| **Age-group specific** |  |  |  |
| LLM |  |  |  |
| 13-29 | 49.6% | 94.2% | 0.84 |
| 30-44 | 45.2% | 97.2% | 0.85 |
| 45+ | 46.3% | 98.7% | 0.88 |
| XGBoost |  |  |  |
| 13-29 | 88.0% | 62.1% | 0.84 |
| 30- | 70.6% | 82.3% | 0.84 |
| 45+ | 62.2% | 93.3% | 0.89 |
| LASSO |  |  |  |
| 13-29 | 89.5% | 54.8% | 0.82 |
| 30- | 75.6% | 74.8% | 0.82 |
| 45+ | 58.2% | 92.7% | 0.87 |
| **Race-specific** |  |  |  |
| **LLM** |  |  |  |
| Non-Hispanic White | 37.8% | 99.3% | 0.86 |
| Non-Hispanic Black | 51.9% | 95.4% | 0.86 |
| Hispanic | 46.4% | 97.9% | 0.86 |
| XGBoost |  |  |  |
| Non-Hispanic White | 50.5% | 95.8% | 0.87 |
| Non-Hispanic Black | 82.7% | 72.8% | 0.86 |
| Hispanic | 62.5% | 88.7% | 0.85 |
| **LASSO** |  |  |  |
| Non-Hispanic White | 44.1% | 95.8% | 0.86 |
| Non-Hispanic Black | 84.7% | 65.8% | 0.84 |
| Hispanic | 65.2% | 84.6% | 0.83 |

Figure 2 depicts the “text area” focused by the GatorTron model, using the LIME package, for three representative cases, highlighting the narrative features that most strongly influence the GatorTron LLM’s assessment of HIV risk. Subword features associated with higher HIV risk include “addiction”, “substance”, “tobacco”, “infection”, “Black or African American”, “Hispanic”, “ibuprofen”, “sodium chloride”, “viral”, and “parasitic”. Subword features associated with lower HIV risk include “female”, “analgesics”, “pain”, “diazepam”, “ischemic heart disease”, “diabetes”, and “gender”.

**Figure 2.**
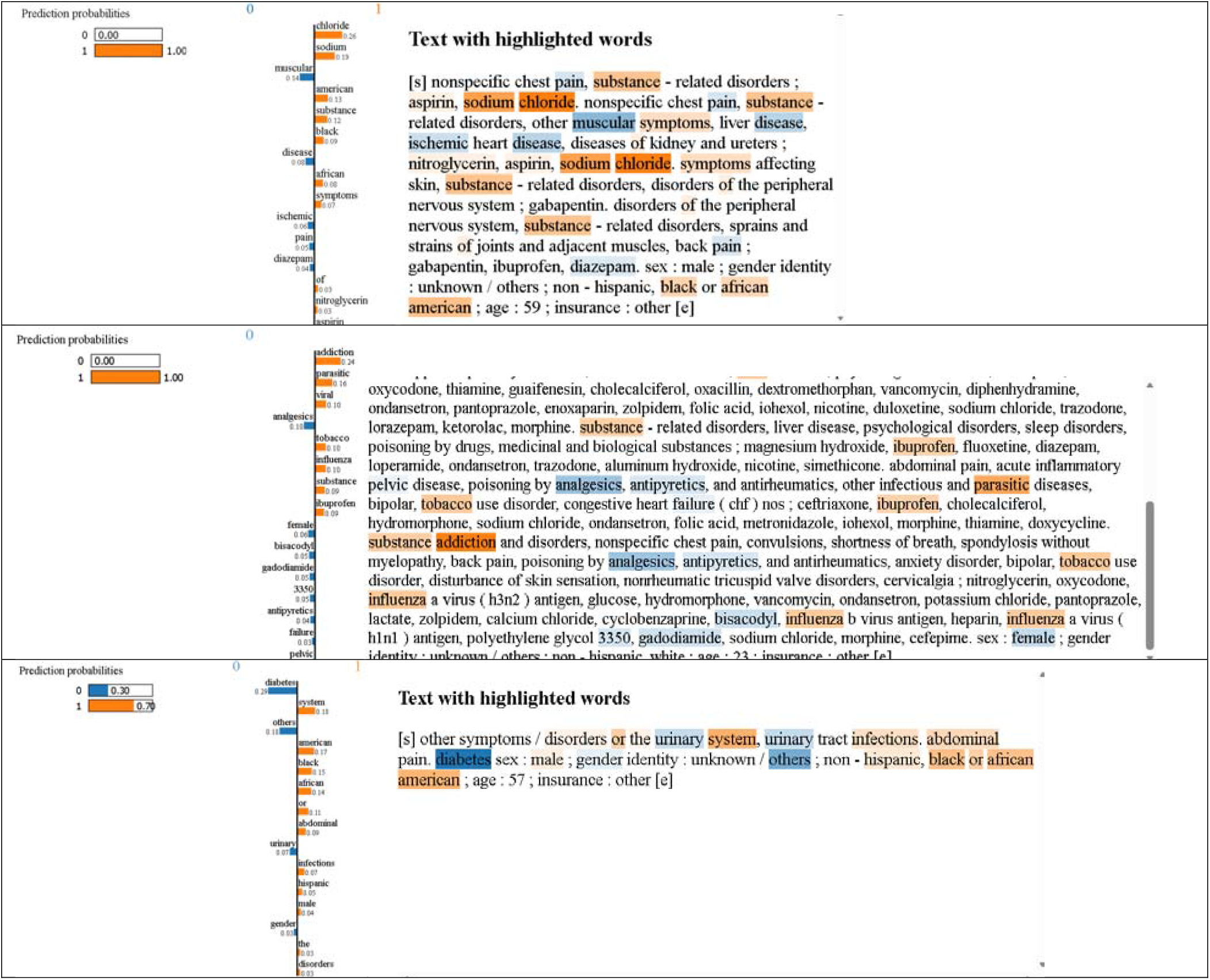
Visualization of feature importance highlighted by LLM GatorTron from three representative cases

The top 20 important features identified by the traditional machine learning models are presented as odds ratios (ORs) with 95% confidence intervals (CIs) for the LASSO model (Table 4) and as mean SHAP values for the XGBoost model (Figure 3). The important features largely overlapped between the two models and included demographic factors, the number of emergency department visits, and Phecodes representing several chronic conditions. Being male (OR = 3.04, 95% CI: 2.47–3.74) and non-Hispanic Black (OR = 4.61, 95% CI: 3.69–5.78) were the two strongest risk factors. In contrast, diagnoses or screenings for several chronic conditions appeared to be protective factors, although some associations were not statistically significant.

**Table 4.**
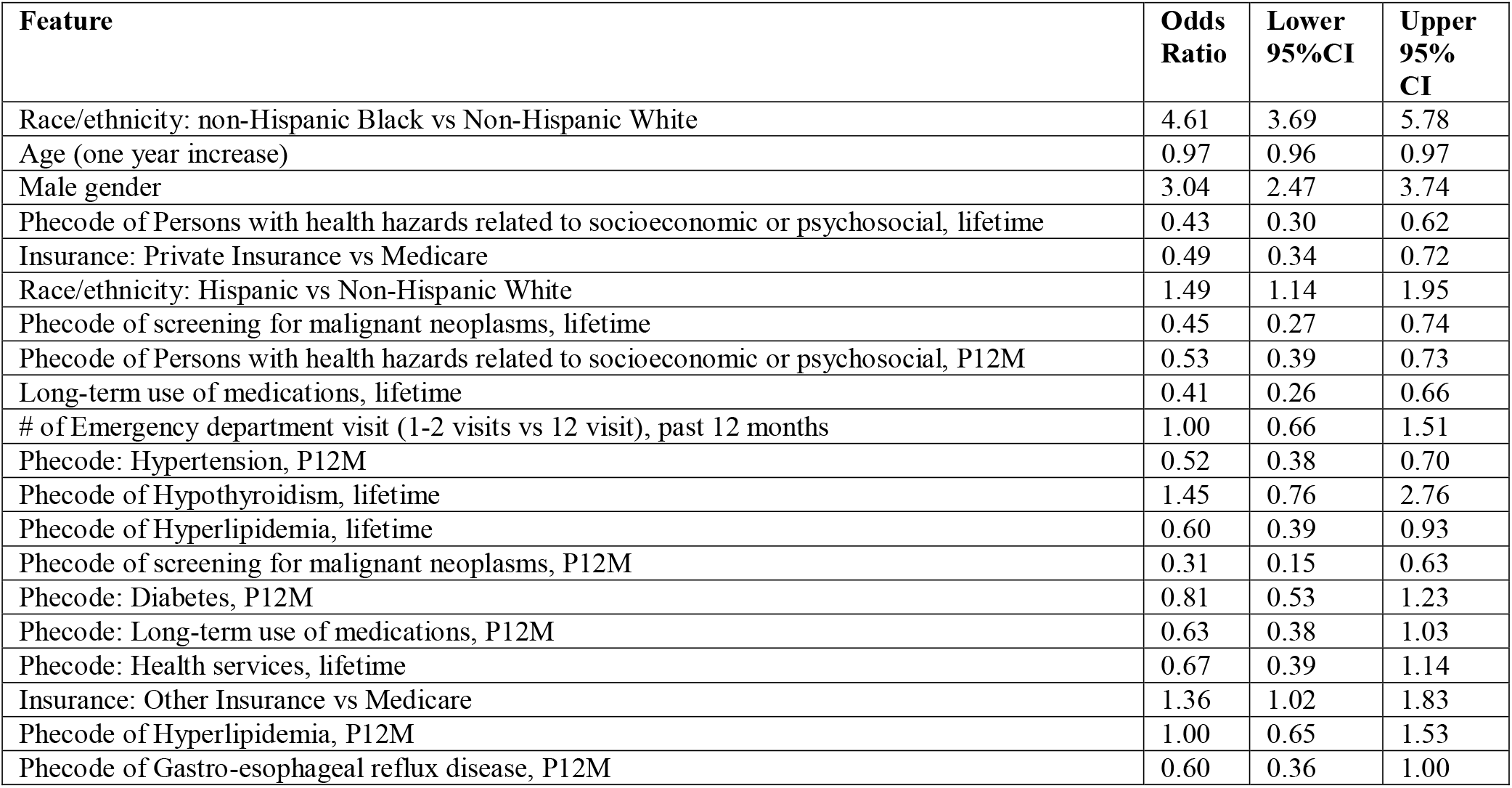
Odds ratio and 95% confidence interval (CI) for the top 20 features from the LASSO model.

**Figure 3.**
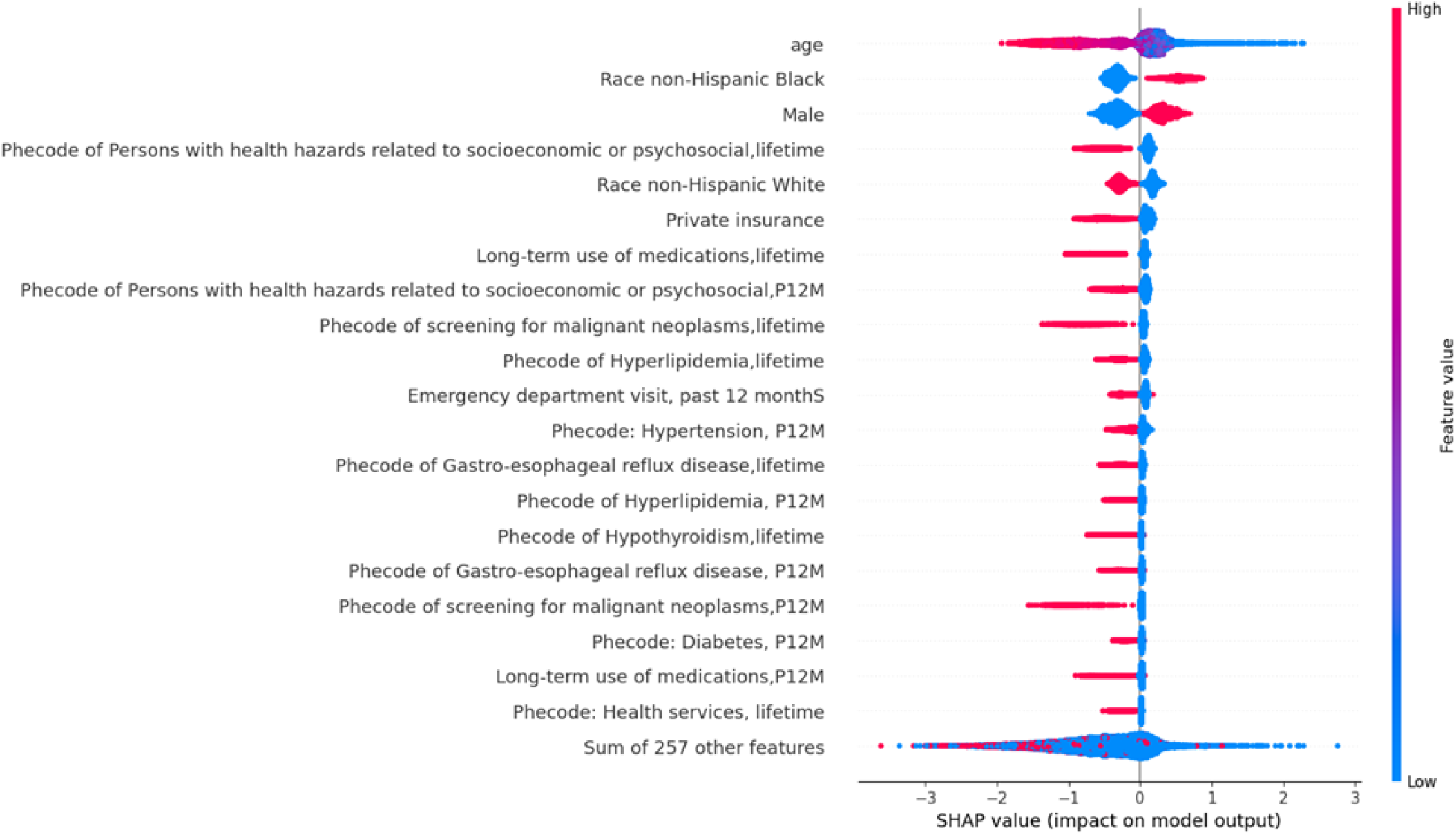
Top 20 Features from XGBoost with the highest mean SHAP values

## Discussion

By converting structured EHR data into narrative text, we showed that LLMs can effectively identify people at HIV risk using EHRs. Our clinical LLM, GatorTron, achieved strong discrimination (AUC = 0.88), comparable to traditional ML methods but with less preprocessing and feature engineering. Both approaches demonstrated model fairness, with consistent performance across demographic groups and interpretable predictors.

Incorporating the four socio-demographic variables (i.e., age, race/ethnicity, sex, and insurance) into the LLM that considered diagnosis data significantly improved model performance. This finding highlights the key role of demographic factors in HIV risk prediction, supported by their strong influence in feature attribution, and is consistent with past HIV risk modeling studies [4, 5, 7, 11, 18]. None of these socio-demographic factors has direct causal effects on HIV acquisition; they enhance prediction performance possibly because they serve as proxies for underlying social and behavioral determinants of health that are not well captured in EHR data, such as access to care, socioeconomic status, stigma, and network-level HIV risk. Importantly, despite their contribution to predictive power, our results showed that the model maintained consistent performance across demographic groups, suggesting that the inclusion of these variables enhanced prediction accuracy without introducing systematic bias.

Including medication information in the LLM, represented by medication active ingredient transformed from RxNorm codes, did not further improve model performance. This differs slightly from our prior study on heart failure risk prediction among cancer patients using the same LLM, where medication data further improved model performance [20]. The difference may be explained by the differences in disease prognosis and treatment responses. Some medications can remarkably influence heart failure risk, but in HIV prediction, medication prescriptions largely align with documented diagnoses and provide limited additional information. To our knowledge, no prior studies have examined the impact of medication data on HIV risk prediction, limiting direct comparison.

Model interpretability analyses revealed broadly consistent findings across approaches. Demographic variables such as male sex and non-Hispanic Black race emerged as strong predictors of HIV risk across all models, and narrative features related to substance use and infection were highlighted by the LLM. These predictors align with known epidemiologic risk factors for HIV and prior HIV risk modeling studies [5, 7, 18, 30, 31]. In contrast, features associated with chronic disease management, such as diabetes and ischemic heart disease, were linked to lower predicted HIV risk in both the LLM and traditional ML models. This pattern is consistent with our prior HIV risk model in women [11] and may reflect greater healthcare engagement among patients with chronic conditions, which is associated with lower HIV vulnerability. Of note, LIME provides local feature attributions that help approximate model behavior, and these outputs do not represent clinical reasoning.

Future studies could explore broader factors related to HIV. First, both community- and individual-level social and behavioral determinants of health (SDOH) can be incorporated into the narrative to improve LLM HIV prediction. Patient ZIP codes could be linked to external datasets to capture neighborhood-level HIV risk factors and SDOH [11], and clinical notes could provide additional individual-level SDOH, such as stigma [32, 33], risky sexual activities. Second, future studies should assess the perceptions of both providers and patients regarding LLM-generated predictions, which are important for building trust and guiding clinical implementation. Third, future studies should investigate domain-specific GPT-based generative LLMs, including their capacity to reason and generate natural language explanations, which may facilitate interactive and interpretable predictions to support clinical decision-making and patient communication.

This study has several limitations. The findings have not been externally validated, and replication in other populations and healthcare systems is needed to confirm performance. Moreover, the prediction was based on EHR data, which is inherently subject to missing or incomplete data and focused on people with access to care, potentially limiting generalizability. Due to the 512-token limit, 7.8% of records were truncated, and the impact of this truncation on model performance remains uncertain. Fairness was assessed by comparing AUC across demographic subgroups; however, this approach captures only one aspect of performance parity and does not fully evaluate other dimensions of fairness.

## Conclusion

Our LLM-based solution, GatorTron, effectively predicts HIV risk from structured EHR data transformed into narrative text, achieving the best F1-score and with minimal preprocessing. Using a local LLM also preserves data privacy by keeping sensitive information within the secure hospital environment. These findings highlight the potential of clinical LLMs to support accurate HIV risk prediction and enhance the delivery of HIV prevention services.

## Data Availability

Data used in the current analysis can be requested by submitting an application to OneFlorida+ https://onefl.net/

## Acknowledgements

This work was supported by the National Institute on Allergy and Infectious Diseases R01AI172875 (MPI Prosperi/Bian), National Institute on Mental Health R34MH135768 (MPI Liu/Cho), and R21MH137736 (MPI Naar/He). Research reported in this publication was supported in part by the OneFlorida Clinical Data Network, funded by the Patient-Centered Outcomes Research Institute (PCORI) numbers CDRN-1501-26692 and RI-CRN-2020-005; in part by PCORI Method Award ME-2023C3-35934, in part by the OneFlorida Cancer Control Alliance, funded by the Florida Department of Health’s James and Esther King Biomedical Research Program number 4KB16; and in part by the University of Florida Clinical and Translational Science Institute and its Clinical and Translational Science Award (CTSA) hub partner, Florida State University, which are supported in part by three CTSA Program grants awarded on August 15, 2015, and renewed for five years on July 2, 2019, by the National Center for Advancing Translational Sciences of the National Institutes of Health.

The UF-FSU CTSA grant numbers are UL1TR001427, KL2TR001429 and TL1TR001428. The content is solely the responsibility of the authors and does not necessarily represent the official views of the Patient-Centered Outcomes Research Institute (PCORI), its Board of Governors or Methodology, the OneFlorida Clinical Research Consortium, the University of Florida’s Clinical and Translational Science Institute, the Florida Department of Health, or the National Institutes of Health.

